# Moving Beyond Duty Hours: Understanding the Contributors to Internal Medicine Resident Workload and Experience

**DOI:** 10.64898/2026.04.08.26349405

**Authors:** Nicholas Bianchina, Cristina Fischer, Karan Rai, Jessica Clawson, Lauren McBeth, Emily Gottenborg, Angela Keniston, Marisha Burden

## Abstract

**Background:** High workload among healthcare workers has increasingly been correlated with poor patient outcomes, inefficient operational and financial outcomes, and burnout. Despite growing literature exploring causes of attending physician workload, there is limited understanding of trainee-specific measures.

**Objective:** We aimed to characterize elements contributing to trainee workload and perceived challenges and satisfiers to the trainee workday as a foundation for better understanding and measuring trainee work experience.

**Methods:** Internal Medicine and Medicine-Pediatrics residents at an academic medical center were invited to participate in focus groups discussing contributors to inpatient workload and work experience between March and April 2024. A qualitative content analysis identified key metrics of trainee workload and work experience, which were then consolidated into overarching domains. A structured, multi-round rating process ranked the perceived relevance of each metric.

**Results:** Twenty residents participated across six focus groups. Analysis of focus groups yielded 297 workload metrics across 28 unique domains. Seventeen domains had metrics identified as highly relevant (median 6-7; IQR ≤ 1) including autonomy, communication, disruptions, task switching, documentation, emotional burden, patient factors, professional fulfillment, rounding, teaming, and work-life balance.

**Conclusions:** Resident physicians highlighted complex interactions between clinical factors, work design, and psychosocial dynamics that contribute to their sense of workload. This creates opportunities to develop unique measures of workload to understand the trainee experience better. Further studies are needed to capture the generalizability of these findings and the relationship between these workload domains and patient, organizational, and trainee outcomes with the aim of implementing evidence-based work design.

## Introduction

Despite extensive clinical guidelines and best practices, there are few evidence-based practices to guide how inpatient physician work should be structured or measured.^1^ This gap persists despite research demonstrating that both workload and work environment directly influence outcomes ^1,2^ – including patient mortality, failure to rescue, physician burnout and moral injury, and even organizational profitability.^3,4^ Lacking robust guidance, existing approaches to workload assessment often rely on readily available metrics such as work relative value units (wRVU), patient census, and duty hours. While such metrics are practical and carry merit, they do not offer a comprehensive capture of the multidimensional nature of clinical work and its relationship to patient, workforce, and organizational outcomes.^1,2^

Prior work focused on nurses and attending physicians has demonstrated that workload is a multidimensional construct linked to both clinical and organizational outcomes. ^3,5–8^ However, efforts to reform trainee workload have historically focused on duty hour restrictions, implemented in response to concerns regarding fatigue and patient safety.^9^ Although, these reforms addressed work duration, they did not resolve broader questions regarding how trainee workload should be defined, measured, and optimized.^10^ Workload extends beyond time-based metrics to include cognitive load, task switching, team dynamics, quality of the work environment, learning milestones, and professional fulfillment, all of which may influence learning, performance, and well-being. Additionally, while personal drivers of burnout have been extensively studies, limited attention to system-level drivers of workload or the structure of the clinical environment.^9–15^

Building on prior work defining workload and environment for attending physicians and advanced practice clinicians,^2,16–20^ we sought to identify and prioritize metrics that capture the workload and experience of internal medicine trainees in the inpatient setting. Using qualitative methods, we aimed to develop a framework of measurable components of trainee work to support future efforts in evidence-based work design. This work is particularly relevant as evolving care models^21^ and emerging technologies^22,23^ continue to reshape clinical workflows and the educational learning environment.

## Methods

### Project design

We conducted a multi-method study combining focus groups, directed content analysis, and a structured, multi-round ranking process to identify and prioritize components of resident workload and work experience in the inpatient setting (FIGURE 1).^17,24^ Guided by elements of the Chokshi and Mann Process Model for User-Centered Digital Development,^25^ we focused on the *Discover* and *Define* phases to generate and refine measurable elements of clinical work. Additionally, we utilized an interpretative framework of pragmatism, we sought to understand real-world challenges encountered by residents with a goal of designing practical solutions. This project received an exempt, quality improvement determination from the Colorado Multiple Institutional Review Board (COMIRB#24-0303, 2/8/24).

**Figure 1:**
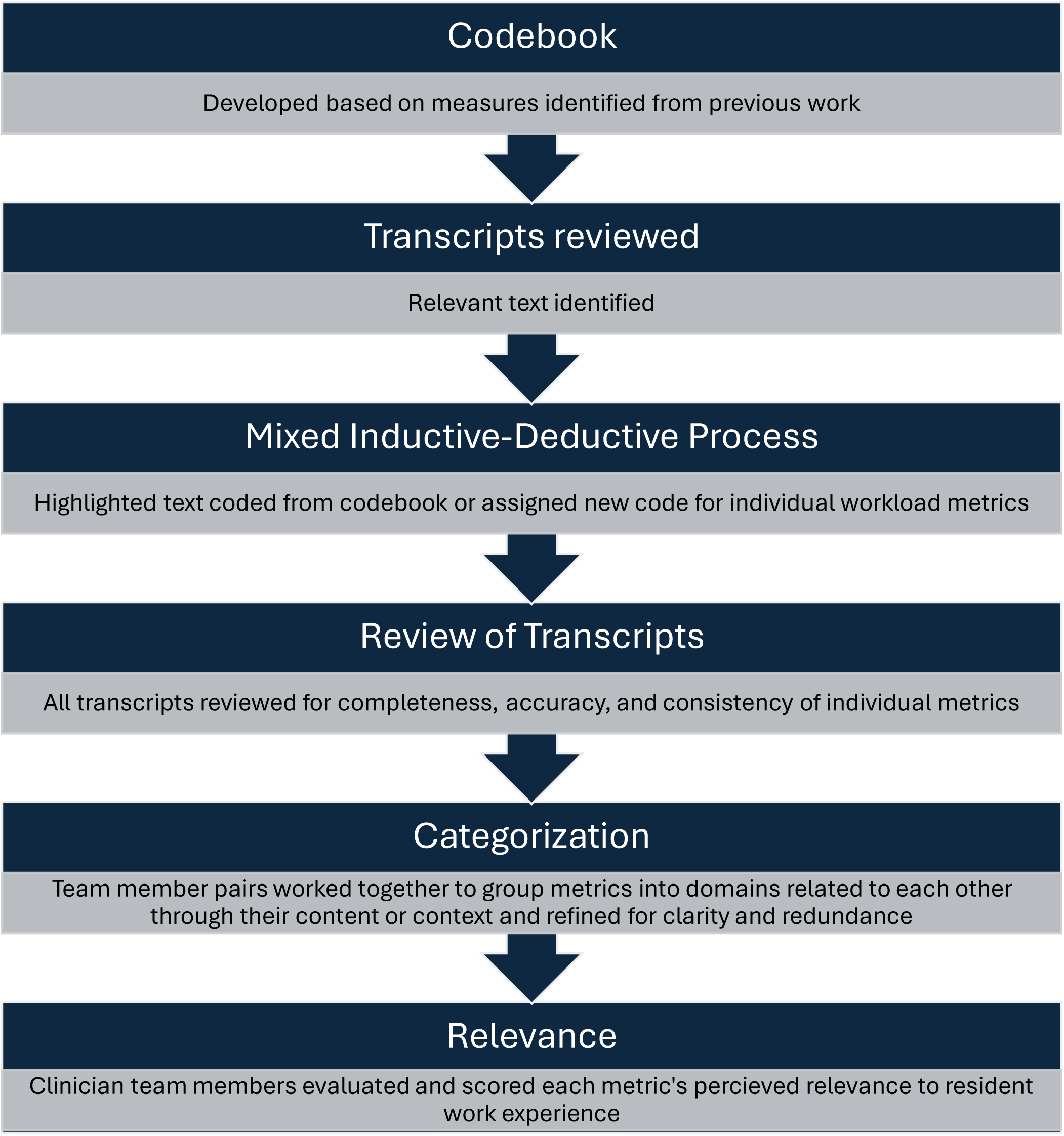
Qualitative Content Analysis Process

### Context and setting

The project was conducted at a 678-bed academic medical center. The residency program has 192 interns and residents.

### Participants and recruitment

Participants included Internal Medicine (IM) and Medicine-Pediatrics (MP) resident physicians in their post-graduate-year (PGY) 1 through 4, with at least six months’ experience in the residency program. The programs are both accredited and in good standing.

We used a convenience sampling approach, inviting all eligible IM and MP residents. Recruitment of eligible residents was conducted via one e-mail sent by a program coordinator outside of the project team, as well as in-person during educational conference time from a non-supervisory resident member of the project team. Preliminary residents were not eligible for participation as the goal was for specific IM resident workload.

The project background and aims were described at the beginning of each focus group, and verbal approval was obtained to participate in the discussion and to collect audio-, visual-, and transcript-based recordings. To protect participant confidentiality, requests to participate in this project did not come from direct supervisors, and participant information was not shared with direct supervisors. Participants received a $10 gift card upon focus group completion, which was supported by internal organizational funds.

### Project team

This project consisted of four IM resident physicians (at the time of project development, focus group completion, and initial data analysis), two hospitalist physicians (one in a hospital medicine leadership role and one who is a member of the IM residency program leadership), and two data analysts and researchers within hospital medicine.

### Focus groups

We conducted six semi-structured virtual focus groups via Zoom Workplace, a video/telephone-based conferencing platform, between March and April 2024. Each session lasted for 45 to 60 minutes and had two to four participants per group. The interview guide included two core questions exploring factors contributing to resident workload and factors influencing work experience, with prompts to elicit specific examples (Supplement). Questions were adapted from findings from prior work.^17^ A brief, anonymous survey was distributed during each session via REDCap^26^ to collect demographic data, including personal and professional attributes (Supplement). One facilitator (i.e., a team member who asked questions and led the group) and one observer (i.e., a team member who completed field notes based on the discussions) were present for each focus group, and team members (AK, CF, JC, KR, LM, NB) alternated through each role. Sessions were transcribed using the Zoom-based transcription function and verified against video recordings (EG).

### Qualitative content analysis

A directed content analysis^27^ of focus group transcripts and recordings was conducted using a mixed inductive-deductive approach. A preliminary codebook was developed using a deductive approach based on previous work.^17^ Transcripts were reviewed, and relevant text reflecting workload or work environment was highlighted and coded either using an existing code from the codebook or, when needed, a new code. All members of the project team jointly reviewed and coded one focus group transcript with discussion as needed to allow for triangulation and to ensure standardization.^28^ The remaining focus groups were subsequently divided amongst team members (AK, CF, JC, KR, LM, MB, NB), with individual review of and coding for each focus group. One team member (EG) reviewed all coding for completeness and accuracy. Codes representing discrete aspects of workload were grouped into metrics, classified as objective (e.g., hours worked) or subjective (e.g., perceived autonomy). Related metrics were then grouped into representative overarching domains (e.g communication). The list of domains was iteratively refined by project team members (CF, JC, KR, NB) to reduce redundancy and exemplar quotes for each domain were selected (FIGURE 1).

### Multi-Round Ranking

Following the directed content analysis, we used a three-round ranking process including five study team members (CF, EG, JC, KR, NB) to prioritize metrics.^24^ Each member of the panel independently and anonymously evaluated and scored each metric for perceived relevance to resident work experience^24^ using a Likert scale from 1 to 7 (1-2 = not relevant, 3-5 = moderately relevant, 6-7 = very relevant). Ratings were summarized with medians, and interquartile ranges (IQR) were calculated to evaluate for consensus among rankings. IQR reflects the spread of responses, with lower values indicating greater agreement. Consensus was reached if the IQR was ≤1. For metrics determined to be at least moderately relevant (i.e., median score ≥ 3) that did not achieve consensus, discussion was conducted amongst the group regarding interpretation, clarity, consistency, and the rating process was repeated until consensus was reached.

### Trustworthiness

The project team included resident physicians, attending physicians who are leaders in resident education and hospital operations, a data specialist, and a PhD in Clinical Sciences to promote diverse perspectives. Field notes and video transcripts were returned to iteratively throughout analysis to ensure that metrics and domains accurately represented the data. The COREQ checklist was utilized in manuscript development.^29^

## Results

Twenty residents participated across six focus groups from March to April 2024. Participants represented all training years, with 15 (75%) in PGY-2 through PGY-4. Among the participants, 9 (45%) trained within the Categorical Track, and 6 (30%), 3 (15%), and 2 (10%) were in the Primary Care, MP, and Hospitalist Tracks, respectively. Fifteen (75%) participants were female (Table 1). At the time of this project, the residency program consisted of 174 residents, including 108 (62.1%) Categorical, 25 (14.4%) Hospitalist, and 24 (13.9%) Primary Care Track residents. Among the residency program, 97 (55.8%) of residents were female, and 16 (9.2%) were MP.

**TABLE 1.** Demographics - Focus Group Participants.

| Characteristic | No. (%) of respondents<br>N=20 |
| --- | --- |
| **Year in Residency** |  |
| PGY-1 | 5 (25%) |
| PGY-2 | 5 (25%) |
| PGY-3 | 9 (45%) |
| PGY-4 | 1 (5%) |
| **Training Track** |  |
| Categorical | 9 (45%) |
| Hospitalist Training Track | 2 (10%) |
| Primary Care Track | 6 (30%) |
| Medicine Pediatrics | 3 (15%) |
| Physician Scientist Training Track | 0 (0%) |
| **Career Plans \*** |  |
| Primary Care | 6 (26%) |
| Hospitalist | 5 (22%) |
| Subspecialty | 12 (52%) |
| Non-Clinical | 0 (0%) |
| Other | 0 (0%) |
| **Identity** |  |
| Cisgender Female/Woman | 15 (75%) |
| Cisgender Male/Man | 5 (25%) |
| Genderqueer/gender non-binary/gender fluid | 0 (0%) |
| Transgender female/woman | 0 (0%) |
| Transgender male/man | 0 (0%) |
| Gender not listed | 0 (0%) |

\*select all that apply

Content analysis yielded 297 unique workload and work experience metrics (Supplement). These were consolidated into 28 unique domains, each representing a group of related metrics and containing 1-38 individual metrics. Following the multi-round ranking process, 17 domains contained metrics rated as highly relevant (median 6-7; IQR ≤ 1). These domains spanned both objective and subjective components of work and included areas such as communication, workflow fragmentation, patient factors, teaming, and time demands. Highly rated within these domains included time to address electronic communications, volume of electronic messages, frequency of interruptions and task switching, number of hours worked, and patient census (Table 2). Exemplar quotes from focus groups representing the various domains are listed in Table 3.

**TABLE 2.**
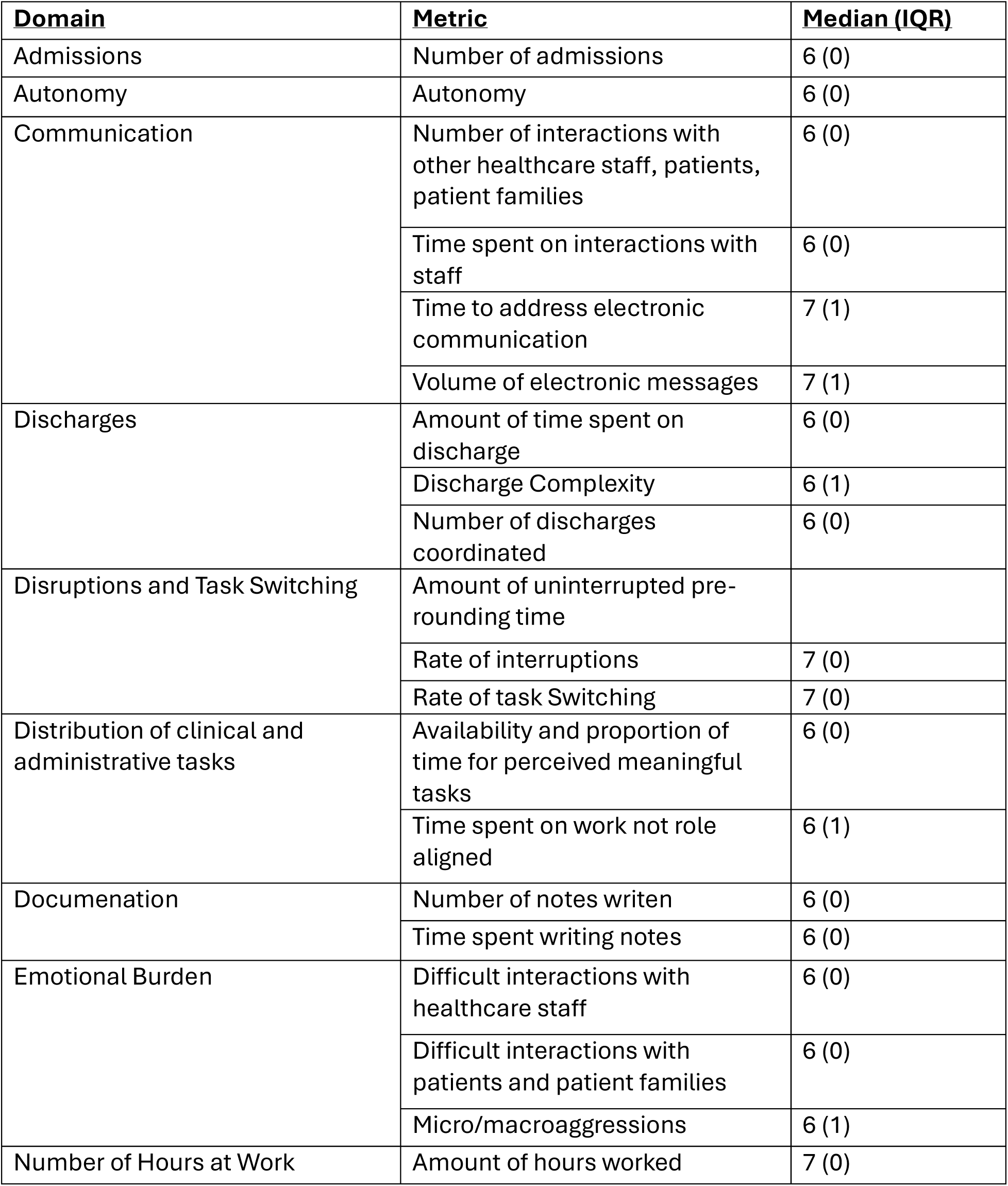
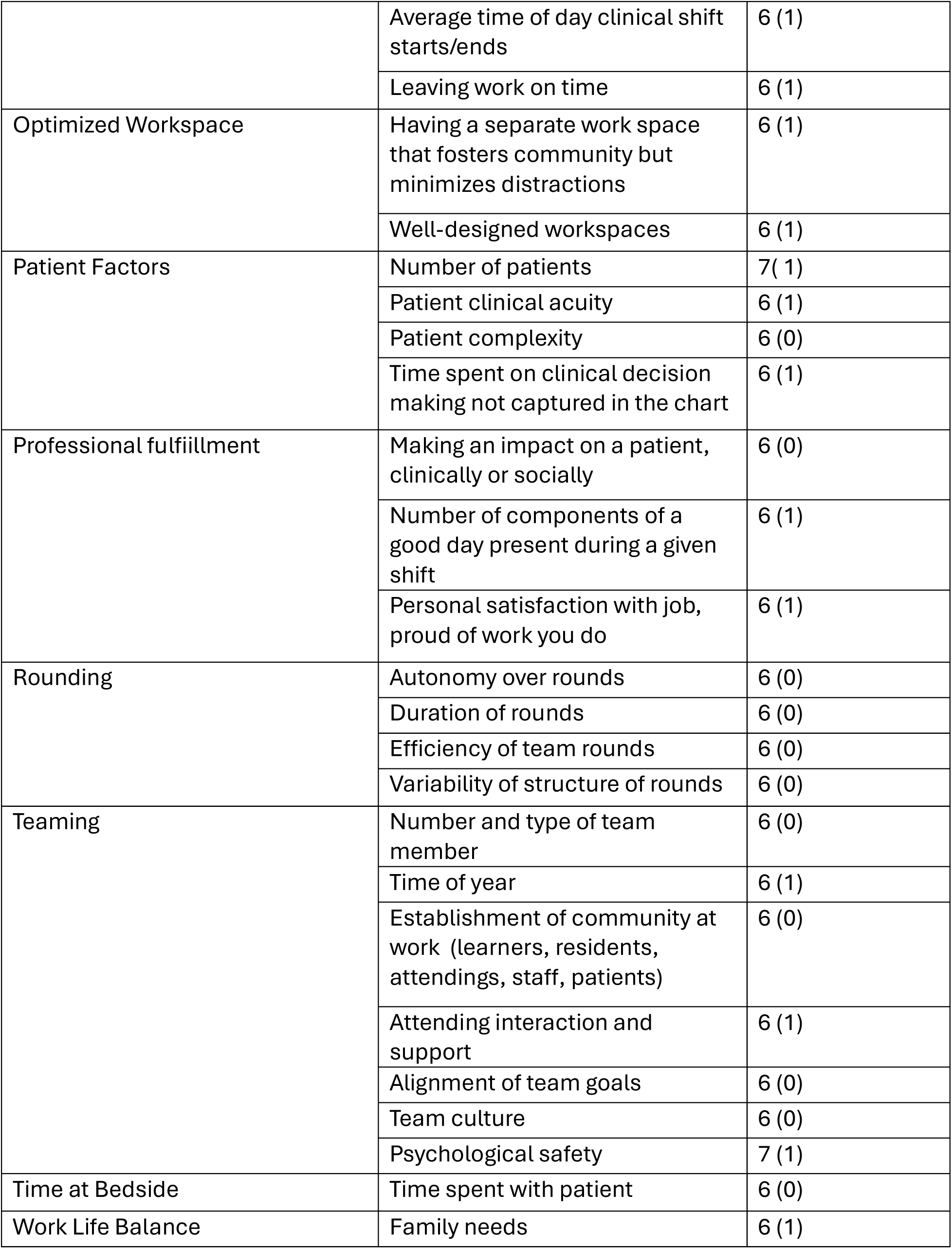
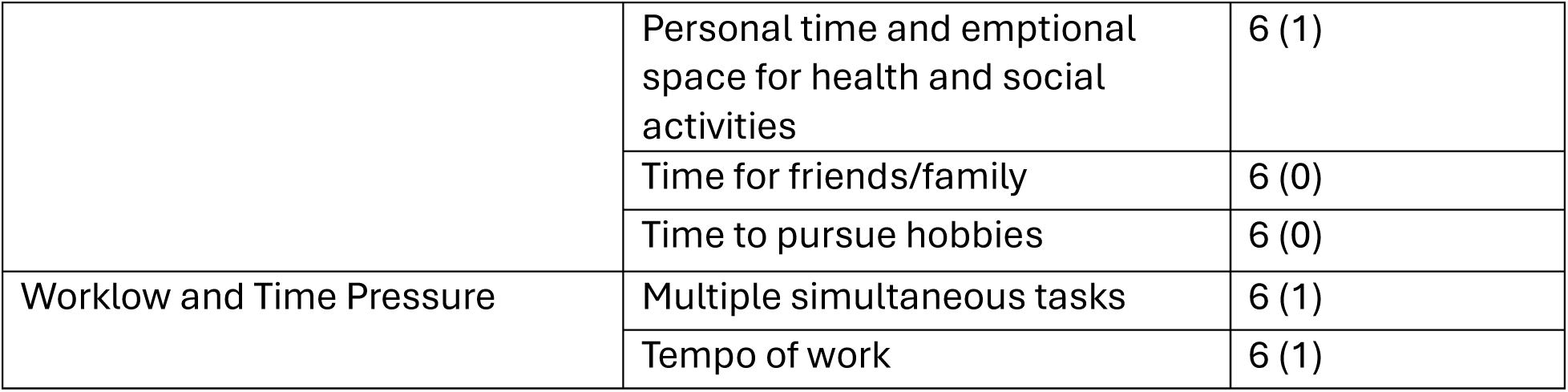
Workload and Work Experience Domains.

**Table 3.**
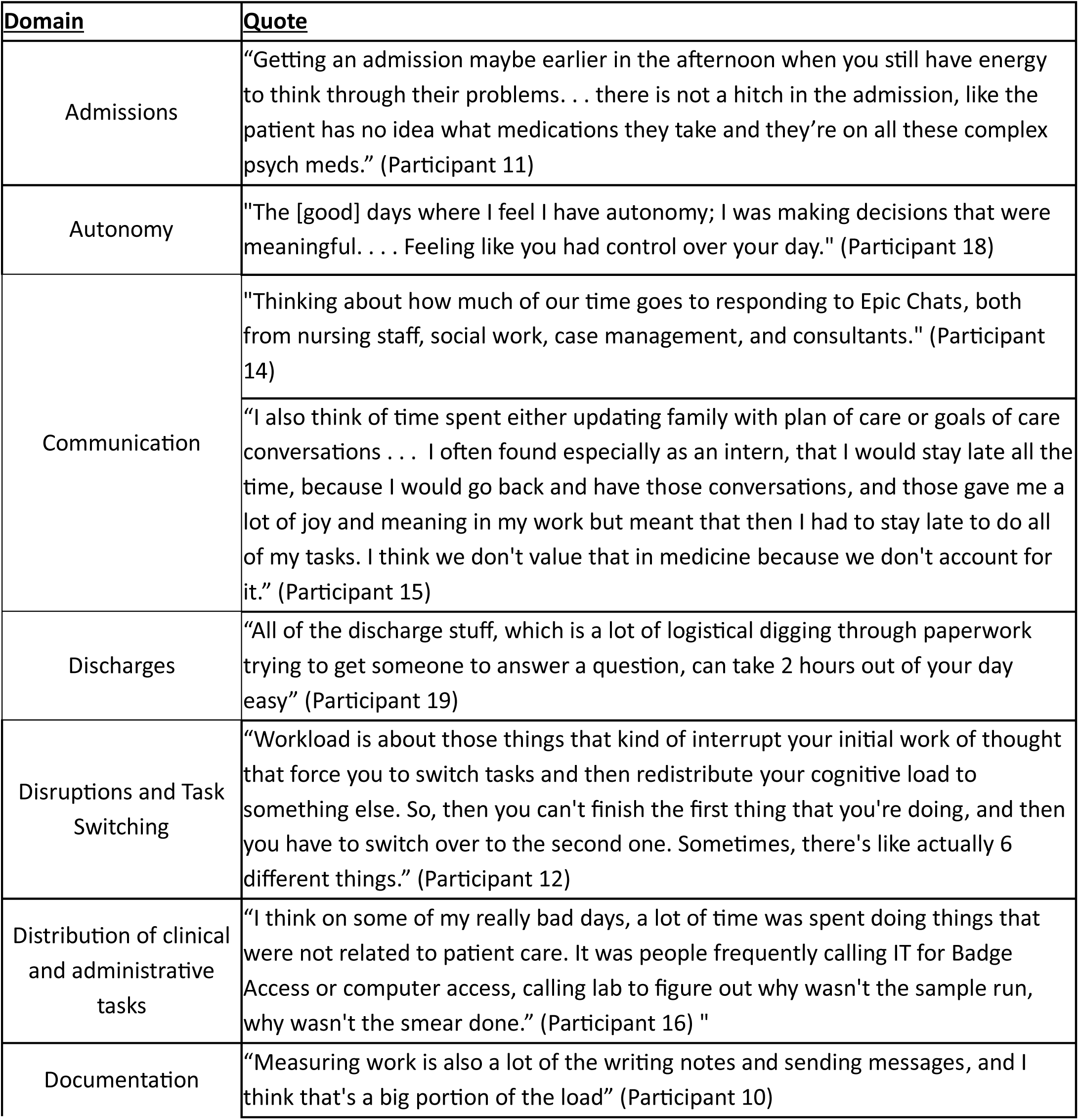
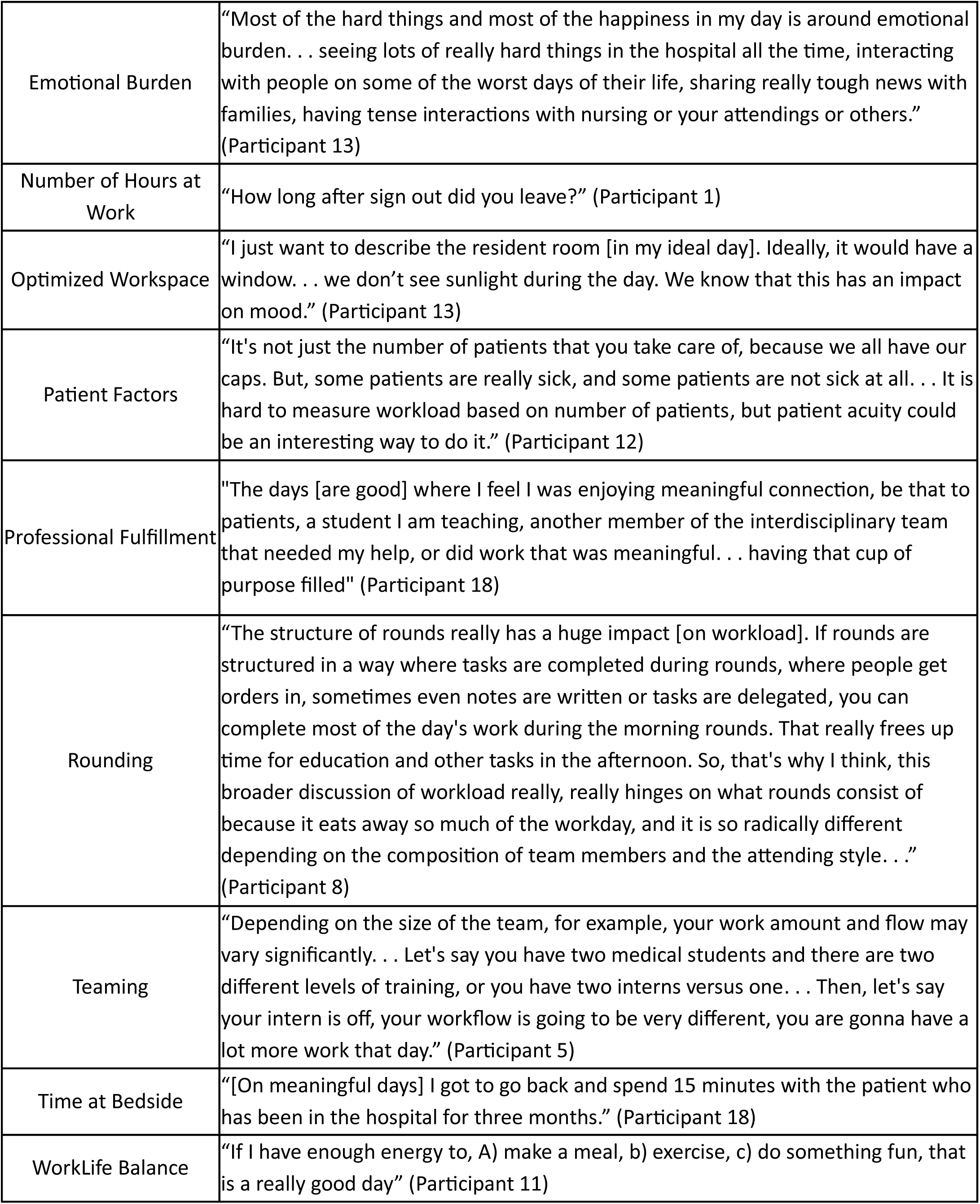
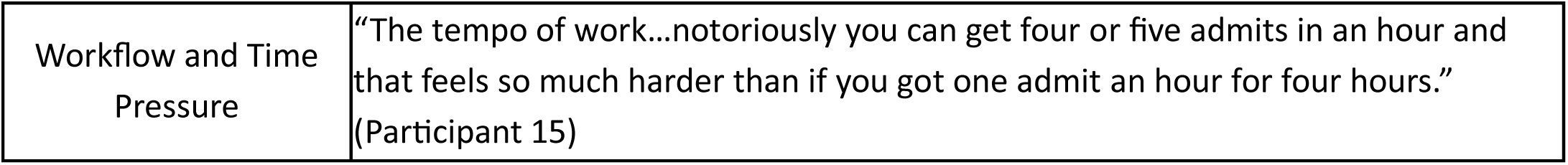
Exemplar Quotes From Each Domain.

| Domain | Quote |
| --- | --- |
| Admissions | "Getting an admission maybe earlier in the afternoon when you still have energy to think through their problems. . . there is not a hitch in the admission, like the patient has no idea what medications they take and they're on all these complex psych meds." (Participant 11) |
| Autonomy | "The [good] days where I feel I have autonomy; I was making decisions that were meaningful. . . . Feeling like you had control over your day." (Participant 18) |
| Communication | "Thinking about how much of our time goes to responding to Epic Chats, both from nursing staff, social work, case management, and consultants." (Participant 14) |
|  | "I also think of time spent either updating family with plan of care or goals of care conversations . . . I often found especially as an intern, that I would stay late all the time, because I would go back and have those conversations, and those gave me a lot of joy and meaning in my work but meant that then I had to stay late to do all of my tasks. I think we don't value that in medicine because we don't account for it." (Participant 15) |
| Discharges | "All of the discharge stuff, which is a lot of logistical digging through paperwork trying to get someone to answer a question, can take 2 hours out of your day easy" (Participant 19) |
| Disruptions and Task Switching | "Workload is about those things that kind of interrupt your initial work of thought that force you to switch tasks and then redistribute your cognitive load to something else. So, then you can't finish the first thing that you're doing, and then you have to switch over to the second one. Sometimes, there's like actually 6 different things." (Participant 12) |
| Distribution of clinical and administrative tasks | "I think on some of my really bad days, a lot of time was spent doing things that were not related to patient care. It was people frequently calling IT for Badge Access or computer access, calling lab to figure out why wasn't the sample run, why wasn't the smear done." (Participant 16) " |
| Documentation | "Measuring work is also a lot of the writing notes and sending messages, and I think that's a big portion of the load" (Participant 10) |
| Emotional Burden | "Most of the hard things and most of the happiness in my day is around emotional burden. . . seeing lots of really hard things in the hospital all the time, interacting with people on some of the worst days of their life, sharing really tough news with families, having tense interactions with nursing or your attendings or others."<br>(Participant 13) |
| Number of Hours at Work | "How long after sign out did you leave?" (Participant 1) |
| Optimized Workspace | "I just want to describe the resident room [in my ideal day]. Ideally, it would have a window. . . we don't see sunlight during the day. We know that this has an impact on mood." (Participant 13) |
| Patient Factors | "It's not just the number of patients that you take care of, because we all have our caps. But, some patients are really sick, and some patients are not sick at all. . . It is hard to measure workload based on number of patients, but patient acuity could be an interesting way to do it." (Participant 12) |
| Professional Fulfillment | "The days [are good] where I feel I was enjoying meaningful connection, be that to patients, a student I am teaching, another member of the interdisciplinary team that needed my help, or did work that was meaningful. . . having that cup of purpose filled" (Participant 18) |
| Rounding | "The structure of rounds really has a huge impact [on workload]. If rounds are structured in a way where tasks are completed during rounds, where people get orders in, sometimes even notes are written or tasks are delegated, you can complete most of the day's work during the morning rounds. That really frees up time for education and other tasks in the afternoon. So, that's why I think, this broader discussion of workload really, really hinges on what rounds consist of because it eats away so much of the workday, and it is so radically different depending on the composition of team members and the attending style. . ."<br>(Participant 8) |
| Teaming | "Depending on the size of the team, for example, your work amount and flow may vary significantly. . . Let's say you have two medical students and there are two different levels of training, or you have two interns versus one. . . Then, let's say your intern is off, your workflow is going to be very different, you are gonna have a lot more work that day." (Participant 5) |
| Time at Bedside | "[On meaningful days] I got to go back and spend 15 minutes with the patient who has been in the hospital for three months." (Participant 18) |
| WorkLife Balance | "If I have enough energy to, A) make a meal, b) exercise, c) do something fun, that is a really good day" (Participant 11) |
| Workflow and Time Pressure | "The tempo of work...notoriously you can get four or five admits in an hour and that feels so much harder than if you got one admit an hour for four hours."<br>(Participant 15) |

## Discussion

This work identifies and prioritizes multi-dimensional components of trainee workload and work experience, extending beyond traditional measures, such as duty hours, patient census.^10,13,15^ We found that workload is shaped by a combination of objective demands (e.g., hours worked, patient volume) and subjective experiences (e.g., autonomy, professional fulfillment, psychological safety), reinforcing the complexity of trainee work and the limitations of characterizing it by time-based metrics alone. This project represents one of the first efforts to define and prioritize metrics of resident workload and work experience within the inpatient setting. ^10,13^ These domains and metrics offer new opportunities for improving trainee work experience. There have been multiple ACGME initiatives to improve overall resident thriving, including integration of well-being into Milestones,^30^ incorporation of burnout and fatigue mitigation in Common Program Requirements^31^ and annual program evaluations, and additional regulatory efforts to ensure that program leaders have sufficient dedicated time to focus on resident experience.^31^ This project lays the groundwork to better understand the factors that may affect resident thriving to help inform future targeted interventions to effectively optimize the multifaceted domains of resident work and to prepare a future thriving physician workforce.

These findings align with prior work^17^ in attending physicians demonstrating the importance of cognitive load, task-switching, EHR interruptions,^32^ electronic communication,^18,33^ discharge planning, complex clinical decision-making, and patient acuity.^34^ This suggests a shared experience among inpatient physicians, regardless of years in practice, and reinforces the core importance of these metrics. Differences in the relative importance of these metrics between trainees and attending physicians requires further investigation. However, some measures of workload previously identified as important by attending physicians^17^ were not named by trainees. These include engagement in quality and safety initiatives, amount of clinical practice time, financial models (e.g., cost of care and savings from readmissions avoided), and intensive care unit days avoided. This may be due to the unique role of trainees that often excludes them from strategic health system quality and safety priorities and medical billing. Our results highlight additional domains particularly relevant to trainees, including rounding, teaming, and psychological safety^17^. While each of these domains has been extensively studied within graduate medical education,^35–37^ their overall effect on resident workload and interplay amongst the other domains remains unclear and presents an opportunity for future work. Lastly, this work is a first step towards evidence-based work design^1^ for IM residency programs – one that integrates empirical data, trainee and educational leader expertise, and rapid, iterative improvement to address workload and work environment. The clinical workplace is rapidly shifting with the emergence of artificial intelligence scribing tools, decision support, and information summarization tools.^38–40^ These technologies and tools are expected to profoundly influence several domains identified in this project, including documentation, communication, cognitive load, and even team dynamics.

This project has several limitations. This was a single-center investigation of two residency specialty programs, which may limit generalizability. When implementing evidence-based work-design, each medical specialty and training location likely will need to tailor approaches for effective implementation in variable local work environments and cultures. Additionally, this project involved residents on the team who ranked the importance of the metrics identified through focus groups. This bias was mitigated by taking an independent rating approach with members across training years and continued blind ranking until agreement was reached. A resident team member was also involved in recruitment, which may lead to selection bias; however, the invitation was open to all members of the training program, and demographic data reflect participation across tracks and years of training.

Future efforts should focus on determining which metrics of workload and work experience are most strongly associated with outcomes of interest, such as patient care quality, trainee well-being, and educational experience, and apply a learning health system framework to continuously evaluate and optimize the resident learning environment^1,41^ as technologies continue to influence it. Additionally, this work sets the foundation for future development of real-time monitoring of various aspects of trainee workload that may allow targeted interventions at the program and individual level. With precision education emerging as a major area of innovation in medical training,^42^ it is critical to ensure that educational interventions are timed with the learner’s cognitive capacity and optimal workload.

## Conclusion

Resident workload is a multi-dimensional construct encompassing both objective demands and subjective experiences. This study defines and prioritizes measurable components of IM trainee work, providing a foundation for future efforts to evaluate and optimize clinical training environments. Moving forward, linking these metrics to outcomes will be essential to inform targeted interventions and support evidence-based redesign of IM residency training. This is particularly important as emerging technologies, including artificial intelligence-enabled documentation and communication tools, reshape clinical workflows.

## Supporting information

Focus Group Guide

## Data Availability

All data produced in the present study are available upon reasonable requested to the authors

## Acknowledgements

None

## Conflicts of Interest

Drs. Burden and Keniston report funding from the Agency for Healthcare Research and Quality, the National Institute for Occupational Health and Safety, University of Colorado Innovations digiSPARK award, and the American Medical Association not related to this work.

Drs. Burden and Keniston contributed to the development of GrittyWork, a digital workforce application, and a registered trademark of the University of Colorado. This application is not currently receiving any financial incentives for its use, and the application was not utilized in this project. Dr. Burden also reports funding from Med-IQ not related to this work.

The authors utilized the ChatGPT language model (version 4o, 5, 5.2) developed by OpenAI for editing original author content to improve readability. All information and materials in the manuscript are original.

Dr. Nicholas Bianchina had access to all the data in the project and takes responsibility for the integrity of the data and the accuracy of the data analysis.

The views expressed in this article are those of the authors and do not necessarily represent the views, policies, or positions of any affiliated institutions or employers. Similarly, concepts presented in the introduction and discussion are intended to illustrate general principles and do not describe or reflect any specific health system or organization.

