## Supplementary material for "Moving Beyond Duty Hours: Understanding the Contributors to Internal Medicine Resident Workload and Experience": Focus Group Guide

**PI:** Marisha Burden

**Moderator - Short presentation on why we are doing this study:**

"Currently there is very little research exploring what contributes to resident workload and how we best measure this. We know that a resident's job is dramatically different from that of an attending clinician and may need different approaches to measure the workload associated with this role. We are exploring contributors to your workload and ideas or challenges in measuring this.

Your participation is strictly voluntary, and all of your responses will be completely anonymous. It is important that what is said here remains confidential so that we can create a safe environment to speak freely without judgment or negative repercussions. We will record our discussion today as we want to be able to capture everything that is said, but we will not be reporting on one specific person or disclosing any information that will identify a person, program, or specific location in the transcript or final reports.

Disclaimer: We are going to divide into breakout groups to have a small group discussion on resident workload. We would like to audio-record these groups to summarize. We will disseminate this information amongst our project team, and potentially use the information as part of future summaries or presentations. We will NOT use the recordings for any other purpose, send them to anyone else, or quote them by name. You can choose not to participate in the small group discussions, and if you decide to participate, you do not have to speak and can leave at any time.

Does everyone agree to these ground rules? Do you have any questions? Are you ready to get started?"

### Focus Group/Interview Guide

**Lead Moderators: In the chat box, introduce yourself. Name and role.**

**Question 1: As we think about how to measure a resident's workday, what would be important to think about measuring?**

> **Prompt**: Consider the current state, where we measure the number of hours worked, or the number of patients that you cared for, what else do you think should we measure?
>
> **Prompt**: We talked a lot about clinical work, are there other things such as educational expectations or scholarly work to explore.
>
> If your group talks about non-clinical work, can flip and state "We talked a lot about non-clinical work, let's talk about patient care workload."
>
> **Prompt**: Think about days that may be challenging? How should we consider measuring that?
>
> **Prompt**: Think about parts of your workday that you enjoy. How should we consider measuring that?

**Question 2:** ***Performed in Round-Robin***

**Describe a perfect day on an inpatient service, where you felt like you were thriving (Defined as 'a state of mental, physical and social functioning to achieve their full potential in work, home and community').**

**Prompt**: What measures in your work environment factor into this?

**Prompt**: What measures outside of work factor into this?

**Prompt**: What measures outside of work factor into this?

------------

**Survey (submitted via survey in REDCap after focus group complete)**

**Demographics:**

1. What is your current training year (PGY-level)?
    a. PGY-1
    b. PGY-2
    c. PGY-3
    d. PGY-4
    e. PGY-5
2. How long have you been in residency ? (months)
    a. 6 – 12
    b. 13 – 18
    c. 19 – 24
    d. 25 – 36
    e. 37+
3. Residency Track:
    a. Hospitalist Training Track
    b. Physician Scientist Training Track
    c. Primary Care Track
    d. Categorical
4. Specialty:
    a. Internal Medicine

   b. Medicine-Pediatrics
   c. Medicine-Psychiatry
   d. Neurology
   e. Ophthalmology
   f. Anesthesia
   g. General Preliminary Year
   h. Other: Free Response

5. Career Plans after Training:
   a. Hospitalist
   b. Primary Care
   c. Subspecialty
   d. (select: Allergy, Cardiology, Endocrinology, Gastroenterology, Hematology/Oncology, Infectious Disease, Nephrology, Pulmonary/Critical Care, Rheumatology, Other)
   e. Non – Clinical
   f. Other
6. Age: Free Response
7. Identify as:
   a. Cisgender female/woman
   b. Cisgender male/man
   c. Genderqueer/gender non-binary/gender fluid
   d. Transgender female/woman
   e. Transgender male/man
   f. Gender not listed here
8. Dependents
   a. Yes: Select number 1-20
   b. No
9. Marital Status:
   a. Single

b. Married
c. Divorced
d. In Relationship, not married
e. Widowed
